# Correlation between Proinflammatory cytokines and severity of COVID-19 within Palestinian Population

**DOI:** 10.1101/2021.08.24.21262535

**Authors:** Walid Basha, Zaher Nazzal, Yousef El-Hamshary, Anwar Odeh, Lama Hijjawi, Mahmoud Doden, Ahmad Musa, Saad Ruzzeh

## Abstract

COVID-19 was characterized by cytokine storm and endothelial dysfunction in severely ill patients. As the severity of the infection was corelated with ethnicity, this study aimed to assess the correlation of proinflammatory cytokine serum level and COVID-19 symptoms within the Palestinian population. In cross-sectional study, serum samples of 27 non-hospitalized patients and 63 hospitalized patients SARS-CoV-2 infected patients, were tested for total antibodies, IL-6, TNF-α, IFN-γ and IL-1β using the ELISA test. Results showed that most common symptoms within patients were Joint pain, cough, and fever (73.3%, 69.7% and 50% respectively). Serum total antibodies (IGs) levels in non-hospitalized patients were higher than hospitalized patients ((44.7 COI and 9.2 COI). TNF-α and IL-6 were lower in non-hospitalized patients compared to hospitalized patients (48±17.9 pg/ml, 193.3±350.5 pg/ml respectively). On the other hand, IFN-γ, in non-hospitalized patients (1±2 IU/ml) was significantly higher than hospitalized patients (0.4±0.26 IU/ml). IL-1β was slightly lower in hospitalized patients (8.8±13.6 pg/ml) compared to non-hospitalized patients (12.5±24.5 pg/ml). Common mild symptoms of COVID-10 were negatively associated with proinflammatory cytokines serum level. In conclusion as it with other populations worldwide, IL-6 and TNF-α are playing a major role in the complications of SARS-CoV-2 infection. Monitoring the two cytokines is crucial for management and treatment of complicated consequence of COVID-19.

## Introduction

Severe acute respiratory syndrome coronavirus-2 (SARS-CoV-2) is a worldwide emerging situation, which was initially reported in December 2019 in Wuhan, China, then affected countries worldwide grew very quickly, and it was declared a pandemic by the WHO [1]. SARS-CoV-2 infection has a heterogenous disease course; it may be asymptomatic in the majority of the cases or mild to severe respiratory infection. More severe cases can be observed such as multi-organ dysfunction syndromes, sepsis and septic shock [2].

Morbidity and mortality in COVID-19 patients has accompanied by the secretion of excessive storm of pro-inflammatory cytokines. induced by the virus. Excessive production of proinflammatory cytokines leads to acute respiratory distress syndrome aggravation and widespread tissue damage resulting in multi-organ failure and death. Targeting cytokines during the management of COVID-19 patients could improve survival rates and reduce mortality. [3]. A classical immunotherapy of convalescent plasma transfusion from recovered patients has been initiated for the neutralization of viremia in terminally ill COVID-19 patients. Due to the limitations of plasma transfusion, researchers are now focusing on developing neutralizing antibodies against virus particles along with immuno-modulation of cytokines like IL-6, Type I interferons (IFNs), and TNF-α that could help in combating the infection [4]. Corelation between the level of pro-inflammatory cytokines were established in many counties and ethnic groups. SARS-CoV-2 infection and hospitalization was varies among race due to genetic variation and chronic disease association [5]. This study aims to investigate the correlation between cytokines level, and specific antibody level with the symptom severity within Palestinian population.

## Material and Methods

### Study design and participants

An observational cross-sectional study was conducted on COVID-19 positive patients. The study was conducted at the Martyrs medical military complex-Corona Hospital-Nablus, which is government-run care facility for managing COVID-19 patients in the North of the West Bank of Palestine. Patients known to have SARS-Cov-2 infection according to the recommendations of the Palestinian Ministry of Health (MOH) on SARS-Cov-2 diagnosis (RT-PCR). Ninety COVID-19 patients were enrolled in the study, sixty-three of them were hospitalized COVID-19 cases with moderate or severe symptoms (Oxygen saturation was less than 92 and need for supplemental Oxygen or has other organ failure) and categorized as the hospitalized patients (HP) and twenty-seven were asymptomatic or with mild symptoms and categorized as non-hospitalized patients (NHP).

### Variables and Data collection

Patient demographic and clinical information, including signs and symptoms, chronic illnesses, and the date of potential exposure to the virus, was gathered from patient clinical records for hospitalized patients and from direct interviews with asymptomatic cases, after the patients or their relatives signed an agreement-consent form to use those data in this study. Venous blood of about 5 ml blood sample were drawn in plane tube from each study participant by a qualified lab technician and transported to the research laboratory at the faculty of Medicine and Health Sciences at An-Najah National University. Serum samples were kept at −80°C in sterile microtubes until the time of ELISA following the manufacturer’s instructions. Samples were taken from all patients before any anti-microbial or anti-inflammatory treatment administrations.

### Total antibodies (IGs) and proinflammatory cytokines measurement

All serum samples were tested for IGs using the ELISA test (Elecsys^®^ Anti-SARS-CoV-2 of the Roche Diagnostics Ltd). According to the manufacturer’s recommendations, samples were considered positive above Cut of index 1 for total antibodies. Proinflammatory cytokines serum concentration was tested by R&D system-Quantikine ELISA for IL-6 and TNF-α and DRG system ELISA for IFN-γ and IL-1β following the manufacture instructions.

### Analysis and Ethical consideration

All statistical analyses were done with IBM SPSS Statistics for Windows, Version 20.0 (IBM Corp., Armonk, NY: IBM Corp). Continuous variables were expressed as mean ± standard deviation (SD). Counts and percentages described categorical variables. The Institutional Review Board (IRB) of An-Najah National University and the Ministry of Health Research Committee approved the study. Informed consent was obtained from each patient involved in this study, or from a member of the patient family. No identifying data were collected during the study and the data were to be only available to the research team.

## Results

### Background characteristics

Age of the study population ranged between 15 and 90 years old with average 57.5 years old. Sample include 56 (62.2%) males and 31 (34.4%) females. The majority were resident in cities (62.2%) and non-smokers (84.4%). Of them, 73.3% were suffering from chronic disease, 51% with diabetes and 61% with hypertension. Previous yearly infection influenza comprised 32.2% of the sample and only 7.8% received annual influenza vaccine.

### Symptoms associated with COVID-19

The symptoms of COVID-19 infection were varying in the incidence among patients. Joint pain was the most encountered symptom, which constitutes approximately 73.3% followed by cough; 69.7% and fever; 50 %. Other symptoms patients experience but at low incidence were weakness; 26.7%, headache; 25.6%, shortness of breath; 23.3%, chest pain; 22.2%, muscle pain; 20%, diarrhoea; 20%, nausea; 17.8%, losing sense of smell; 18.9%, losing sense of taste; 16.7% and sore throat; 14.6%. (Figure 1)

**Figure 1:**
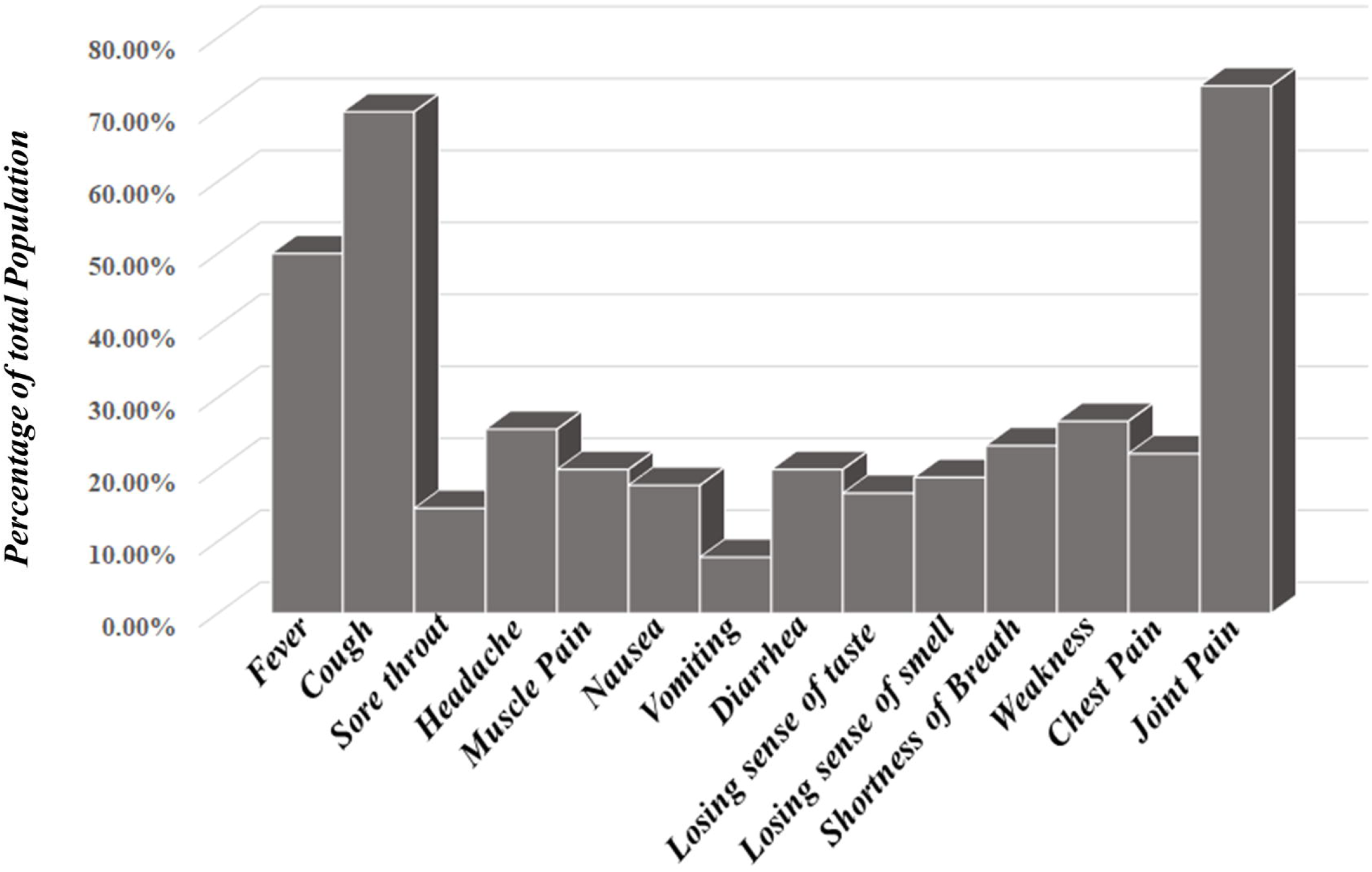
Common symptoms distribution within study population. Columns represent the percentage of patients experience symptoms during COID-19.

### Correlation between severity of COVID-19 and total IGs and cytokines serum level

Serum IGs levels in NHP (44.7±56.6 COI) were significantly higher than HP (9.2 ±14.7 COI), (p=0.012). TNF-α and IL-6 were significantly (p<0.001) lower in NHP compared to HP (48±17.9 pg/ml, 193.3±350.5 pg/ml respectively). In the contrary, IFN-γ, in NHP (1±2 IU/ml) was significantly higher than HP (0.4±0.26 IU/ml) (P=0.001). With no significant difference (p=0.827), IL-1β was slightly lower in HP (8.8±13.6 pg/ml) compared to NHP (12.5±24.5 pg/ml). (Table-1)

**Table 1:** Total IGs, TNF-α, IL6, IFN-γ, IL-1β levels between hospitalized and non-hospitalized COVID-19 patients. \**Mann-Whitney U test*

|  | Hospitalized<br>(n=63) | Non-Hospitalized<br>(n=27) | P-value* |
| --- | --- | --- | --- |
| Total IGs (COI) | 9.2 (± 14.7) | 44.7 (± 56.6) | 0.012 |
| TNF-α (pg/ml) | 193.3 (± 350.5) | 48.0 (± 17.9) | <0.001 |
| IL-6 (pg/ml) | 18.2 (± 34.6) | 4.2 (± 0.88) | <0.001 |
| IFN-γ (IU/ml) | 0.4(± 0.26) | 1.0 (± 2.0) | 0.001 |
| IL-1β (pg/ml) | 8.8 (± 13.6) | 12.5 (± 24.5) | 0.827 |

### Correlation between Cytokine serum level and common symptoms

Cytokine’s level did not show any difference in the absent or presence of nausea, vomiting, diarrhea, shortness of breath, weakness, chest pain and joint pain. Meanwhile the TNF-α concentration was significantly higher in the absent rather than presence of sore throat, headache, muscle pain, losing of taste (Figure 2 A). It was similar with and IL-6 except a significant elevation in the presence of cough (Figure 2 B). On the other hand, IFN-γ were higher in the presence rather than the absent of the symptoms with significant elevation in presence of headache, muscle pain and losing of taste (Figure 2 C). IL-1β level was significantly higher the presence of headache and slightly with muscle pain and the absence of other symptoms (Figure 2 D).

**Figure 2:**
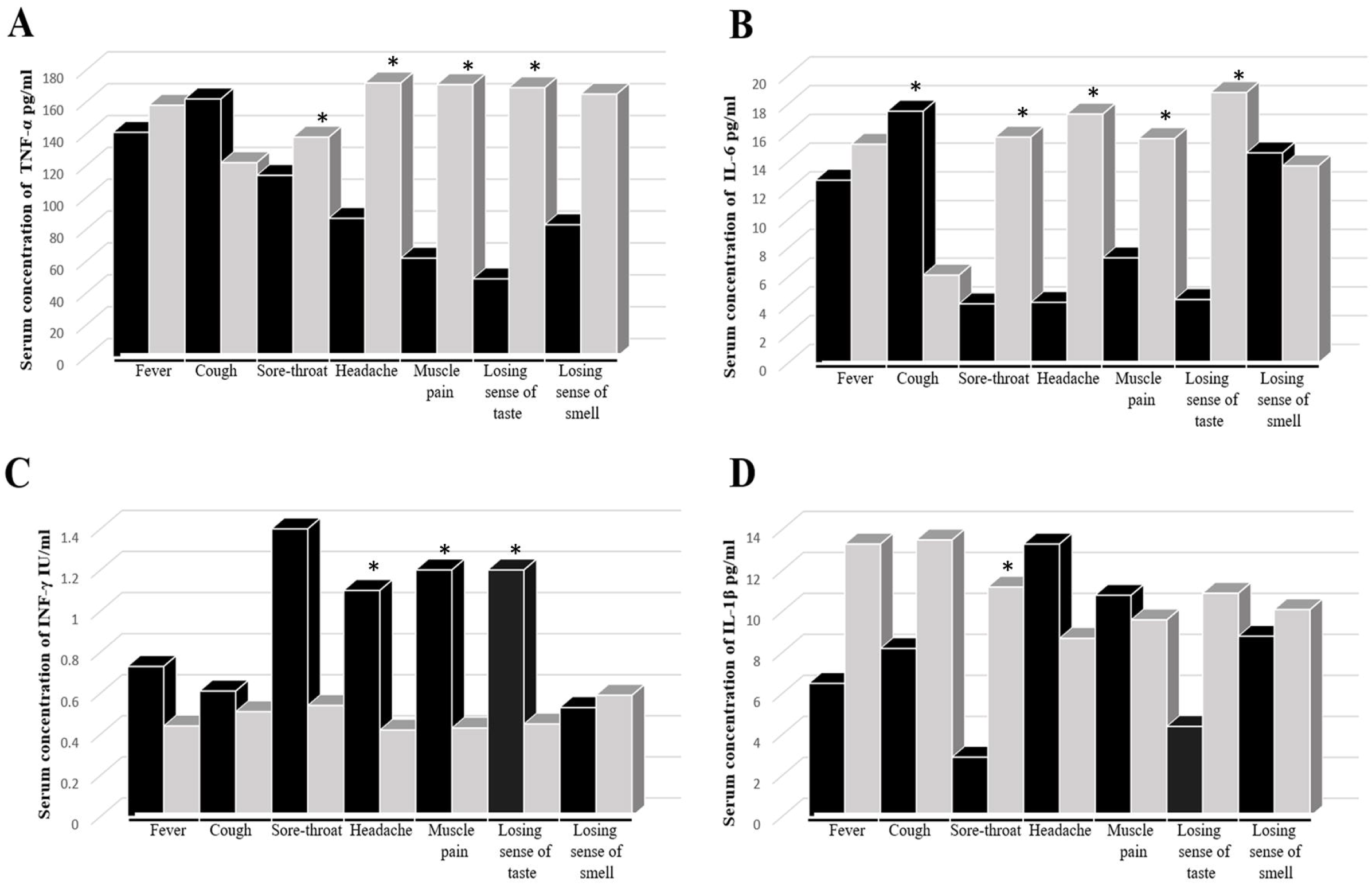
Ccorrelation between the common symptoms and cytokine serum concentrations. Columns represent the serum concentration of TNF-α (A), IL-6 (B), IFN-γ (C) and IL-1β (D) in the presence (Black) and absence (Grey) of common symptoms. \**P. Value <0.05*

## Discussion

Cytokine profiles and immune response in COVID-19 patients were deeply investigated in many countries all over the world, due to the corelation between the severity of the disease and cytokine storm [6]. As in any other viral infection, pro-inflammatory cytokines and chemokines play an important role in immunopathology during viral infections which leading to hyperinflammatory responses in the pathogenesis of COVID-19 disease [7,8]. The incident, severity and the complications of COVID-19 was varied between races and ethnic background [5,9]. This variation refer to the differences in the immune repose and cytokine production between races [10,11] which could be due to allelic distribution and genotype frequency, such as IL-2 and IL-6 [12].

Reports indicated the elevation of serum level of several proinflammatory cytokines, including IL-1β, TNF-α, IL-6 and T-cell cytokine IFN-γ in COVID-19 infection in general despite the severity of the disease [8,13,14]. The correlation with the severity of COVID-19 infection was confirmed with IL-6 [15,16] but with variable results for other cytokines[17–19]. This variation in results lead us to study the relationship of clinical symptom severity with the level of four cytokines within Palestinian COVID-19 patients. Our data showed significant correlation between the severity of COVID-19 and elevation level of IL-6 and TNF-α, with nonsignificant elevation of IFN-γ, with no deference in IL-1β level between HP and NHP. Our study is in close corroboration with De Valle et al who suggested that IL-6 and TNF-α can be independent predictors of the disease severity[19]. This is however in contrast with Chen et al who demonstrated an increase in the IL-6 levels, but the concentration of TNF-α, IL-1, remained unaltered in severely affected patients [17]. The difference in TNF-α concentrations could be due to the race/ethnic background, as well as to the pathological condition and the difference in sampling times. However further research needed for this topic.

The role of antibodies in severity of COVID-19 is less clear. Normally, viral elimination requires cell-mediated immunity, but humoral immunity plays an important role in the elimination of viruses and infected cells such as antibody-dependent cell cytotoxicity (ADCC), opsonization and phagocytosis via innate immune cells. However, two COVID-19 cases of patients with X-linked gamma globulinemia was challenged with antibodies acquired and survived SARS-CoV-2 infection without any sever complications [20]. Some studies have demonstrated and suggested pathogenic role for antibodies in primary infection through enhancement and increased inflammation [21], although this is thought to be not enough to explain the prevalence of severe cases infection [22]. As such, the beneficial, neutral, or harmful role of antibodies in active coronavirus infection remains controversial. Our results didn’t show any correlation with the severity of the infection.

In the contrary with the severity of COVID-19, our results showed that IL-6 and TNF-α were neutral or negatively associated with the common symptoms except for cough, and for IL-6 which was associated with losing since of smell. Previous results suggested a major role of IL-6 and TNF-α smell and taste disfunction [23,24]. This deference may be due to the deference in population were those studies compared serum level of cytokines and chemokines in correlation with SARC-CoV-2 infection with healthy subjects, not correlation with the severity of the disease.

This study has some limitations, including a limited sample size and a lack of time to follow up on patient changes. These factors may have played a role in the lack of a significant association of some of our findings, however they do not change the absence of any correlation between specific clinical severe symptoms and cytokine levels.

## Conclusion

IL-6 and TNF-α have been playing a major role in the complications and severity of SARS-CoV-2 infection within Palestinians like other populations worldwide. But the difference was regarding the relationship of cytokines and common symptoms of infection. Monitoring IL-6 and TNF-α serum level during disease stages is critical to the management and treatment of complicated consequence of COVID-19.

## Data Availability

The data that support the findings of this study are available from the corresponding author, [W. Basha], upon reasonable request.

## Acknowledgment

We thank medical staff at Martyrs medical military complex-Corona Hospital-Nablus for assistance with blood sampling and data collection, and for laboratory technicians at the research lab centre at An-Najah National university for their support in lab work. This work was funded by An-Najah National University.

## References

1 Lai C-C, Shih T-P, Ko W-C, et al. Severe acute respiratory syndrome coronavirus 2 (SARS-CoV-2) and coronavirus disease-2019 (COVID-19): The epidemic and the challenges. Int J Antimicrob Agents 2020;55:105924. doi:10.1016/j.ijantimicag.2020.105924

2 Aminjafari A, Ghasemi S, Robson B, et al. Since January 2020 Elsevier has created a COVID-19 resource centre with free information in English and Mandarin on the novel coronavirus COVID-19. The COVID-19 resource centre is hosted on Elsevier Connect, the company’s public news and information. Brain Behav Immun 2020;S0889-1591:30511-0.

3 Ragab D, Salah Eldin H, Taeimah M, et al. The COVID-19 Cytokine Storm; What We Know So Far. Front Immunol 2020;11:1446. doi:10.3389/fimmu.2020.01446

4 Shah VK, Firmal P, Alam A, et al. Overview of Immune Response During SARS-CoV-2 Infection: Lessons From the Past. Front Immunol 2020;11:1949. doi:10.3389/fimmu.2020.01949

5 Kopel J, Perisetti A, Roghani A, et al. Racial and Gender-Based Differences in COVID-19. Front Public Heal 2020;8:1–8. doi:10.3389/fpubh.2020.00418

6 Akbari H, Tabrizi R, Lankarani KB, et al. The role of cytokine profile and lymphocyte subsets in the severity of coronavirus disease 2019 (COVID-19): A systematic review and meta-analysis. Life Sci 2020;258:118167. doi:10.1016/j.lfs.2020.118167

7 Channappanavar R, Perlman S. Pathogenic human coronavirus infections: causes and consequences of cytokine storm and immunopathology. Semin Immunopathol 2017;39:529–39. doi:10.1007/s00281-017-0629-x

8 Qin C, Zhou L, Hu Z, et al. Dysregulation of Immune Response in Patients With Coronavirus 2019 (COVID-19) in Wuhan, China. Clin Infect Dis an Off Publ Infect Dis Soc Am 2020;71:762–8. doi:10.1093/cid/ciaa248

9 Debnath M, Banerjee M, Berk M. Genetic gateways to COVID-19 infection: Implications for risk, severity, and outcomes. FASEB J Off Publ Fed Am Soc Exp Biol 2020;34:8787–95. doi:10.1096/fj.202001115R

10 Schirmer M, Kumar V, Netea MG, et al. The causes and consequences of variation in human cytokine production in health. Curr Opin Immunol 2018;54:50–8. doi:10.1016/j.coi.2018.05.012

11 Haralambieva IH, Ovsyannikova IG, Kennedy RB, et al. Race and sex-based differences in cytokine immune responses to smallpox vaccine in healthy individuals. Hum Immunol 2013;74:1263–6. doi:10.1016/j.humimm.2013.06.031

12 Cox ED, Hoffmann SC, DiMercurio BS, et al. Cytokine polymorphic analyses indicate ethnic differences in the allelic distribution of interleukin-2 and interleukin-6. Transplantation 2001;72:720–6. doi:10.1097/00007890-200108270-00027

13 Huang C, Wang Y, Li X, et al. Clinical features of patients infected with 2019 novel coronavirus in Wuhan, China. Lancet (London, England) 2020;395:497–506. doi:10.1016/S0140-6736(20)30183-5

14 He R, Lu Z, Zhang L, et al. The clinical course and its correlated immune status in COVID-19 pneumonia. J Clin Virol Off Publ Pan Am Soc Clin Virol 2020;127:104361. doi:10.1016/j.jcv.2020.104361

15 Galván-Román JM, Rodríguez-García SC, Roy-Vallejo E, et al. IL-6 serum levels predict severity and response to tocilizumab in COVID-19: An observational study. J Allergy Clin Immunol 2021;147:72–80.e8. doi:10.1016/j.jaci.2020.09.018

16 Broman N, Rantasärkkä K, Feuth T, et al. IL-6 and other biomarkers as predictors of severity in COVID-19. Ann Med 2021;53:410–2. doi:10.1080/07853890.2020.1840621

17 Chen L, Liu HG, Liu W, et al. [Analysis of clinical features of 29 patients with 2019 novel coronavirus pneumonia]. Zhonghua jie he he hu xi za zhi = Zhonghua jiehe he huxi zazhi = Chinese J Tuberc Respir Dis 2020;43:E005. doi:10.3760/cma.j.issn.1001-0939.2020.0005

18 Zhang J, Walters EH, Tang MLK, et al. Serum cytokine concentrations and asthma persistence to middle age. Allergy. 2020;75:2985–8. doi:10.1111/all.14448

19 Del Valle DM, Kim-Schulze S, Huang H-H, et al. An inflammatory cytokine signature predicts COVID-19 severity and survival. Nat Med 2020;26:1636–43. doi:10.1038/s41591-020-1051-9

20 Soresina A, Moratto D, Chiarini M, et al. Two X-linked agammaglobulinemia patients develop pneumonia as COVID-19 manifestation but recover. Pediatr allergy Immunol Off Publ Eur Soc Pediatr Allergy Immunol 2020;31:565–9. doi:10.1111/pai.13263

21 Liu L, Wei Q, Lin Q, et al. Anti-spike IgG causes severe acute lung injury by skewing macrophage responses during acute SARS-CoV infection. JCI insight 2019;4. doi:10.1172/jci.insight.123158

22 Arvin AM, Fink K, Schmid MA, et al. A perspective on potential antibody-dependent enhancement of SARS-CoV-2. Nature 2020;584:353–63. doi:10.1038/s41586-020-2538-8

23 Cazzolla AP, Lovero R, Lo Muzio L, et al. Taste and Smell Disorders in COVID-19 Patients: Role of Interleukin-6. ACS Chem Neurosci 2020;11:2774–81. doi:10.1021/acschemneuro.0c00447

24 Torabi A, Mohammadbagheri E, Akbari Dilmaghani N, et al. Proinflammatory Cytokines in the Olfactory Mucosa Result in COVID-19 Induced Anosmia. ACS Chem Neurosci 2020;11:1909–13. doi:10.1021/acschemneuro.0c00249

